# A diagnostic protein assay for differentiating follicular thyroid adenoma and carcinoma

**DOI:** 10.1101/2024.09.26.24314403

**Authors:** Yaoting Sun, He Wang, Lu Li, Jianbiao Wang, Wanyuan Chen, Li Peng, Pingping Hu, Jing Yu, Xue Cai, Nan Yao, Yan Zhou, Jiatong Wang, Yingrui Wang, Liqin Qian, Weigang Ge, Mengni Chen, Feng Yang, Zhiqiang Gui, Wei Sun, Zhihong Wang, Minghua Ge, Yi He, Guangzhi Wang, Yongfu Zhao, Huanjie Chen, Xiaohong Wu, Yuxin Du, Wenjun Wei, Fan Wu, Dingcun Luo, Xiangfeng Lin, Haitao Zheng, Xin Zhu, Bei Wei, Jiafei Shen, Jincao Yao, Zhennan Yuan, Tong Liu, Jun Pan, Yifeng Zhang, Yangfan Lv, Qiaonan Guo, Qijun Wu, Tingting Gong, Ting Chen, Shu Zheng, Jingqiang Zhu, Hanqing Liu, Chuang Chen, Hong Han, Sathiyamoorthy Selvarajan, Mingzhao Xing, Kennichi Kakudo, Erik K. Alexander, Yijun Wu, Yu Wang, Dong Xu, Hao Zhang, Xiu Nie, Oi Lian Kon, N. Gopalakrishna Iyer, Zhiyan Liu, Yi Zhu, Haixia Guan, Tiannan Guo, We-TEC Investigators

## Abstract

Differentiating follicular thyroid adenoma (FTA) from carcinoma (FTC) remains challenging due to similar histological features separate from invasion. In this study, we aim to develop and validate DNA and protein-based classifiers for FTA/FTC differentiation. We collected 2443 samples from 1568 patients across 24 centers and applied next-generation sequencing, as well as discovery and targeted proteomics. Machine learning models were developed and compared utilizing DNA and/or protein features. The discovery protein-based model (AUC 0.899) outperformed the gene-based model (AUC 0.670). Consequently, we generated a protein-based model with targeted mass spectrometry and further validated it in three independent testing sets. The 24-protein-based model achieved high performance in the retrospective sets (AUC 0.871 and 0.853) and the prospective fine-needle aspiration biopsies (AUC 0.781). The classifier notably illustrated a 95.7% negative predictive value for ruling out malignant nodules. This study offers a promising protein-based approach for differential diagnosis of FTA and FTC.

**Highlights:**

- Genetic and proteomic profiling of follicular thyroid tumors from 1568 patients across 24 centers
- AI model based on proteins (AUC 0.899) outperformed that based on gene mutations (AUC 0.670) for differentiation of FTA and FTC
- Validation of the protein model in two retrospective cohorts (AUCs: 0.871, 0.853) and a prospective (AUC 0.781)
- The protein model has 95.7% negative predictive value for ruling out malignant nodules

## Introduction

The incidence of thyroid nodules and thyroid cancer has continuously increased over the past decades^1^. Although ultrasonography and ultrasound-guided fine needle aspiration (FNA) improve the distinction between benign and malignant nodules, approximately 10-30% of nodules still cannot be definitively diagnosed by FNA and require surgical diagnosis^2^. These indeterminate thyroid nodules (ITN) are mainly composed of follicular thyroid adenoma (FTA) and follicular thyroid carcinoma (FTC)^3^.

The incidence of benign FTA is approximately five times higher than that of invasive FTC in surgical specimens^4^. Among all FTC patients, 7-23% will have distant metastases at diagnosis or during follow-up^5,6^, and 11-39% will experience recurrence^7,8^. The 10-year disease-specific mortality rate for invasive FTC is 15-28%. Therefore, it is crucial to accurately differentiate FTC from benign conditions to ensure appropriate clinical management and prognosis.

The differential diagnosis between FTA and FTC is one of the most challenging aspects of thyroid pathology due to their subtle differences. Currently, the standard for stratification is based on capsular and vascular invasion^9^. FTA and FTC cannot be distinguished preoperatively because capsular invasion cannot be assessed by cytology, ultrasound, or clinical features. The only way to differentiate them is through diagnostic surgery followed by histopathological examination by experienced pathologists. However, even in postoperative histopathology, FTC and FTA can be challenging to distinguish as FTC often closely resembles FTA microscopically. Capsular invasion must be carefully inspected under the microscope with serial sections to make a judgment. Sometimes, due to insufficient sampling, pathologists are unable to examine the entire capsule in the tissue specimen, making it difficult to provide a definitive diagnosis.

Nucleic acid-based molecular testing has been developed and validated for assistance with ITN and has achieved high negative predictive values (>95%)^3,10,11^. However, genomic and transcriptomic signatures distinguishing FTA from FTC have not yet been identified. *RAS* mutations and PAX8/PPARγ rearrangements are common alterations in follicular neoplasms but can be detected in both, so individual gene alterations cannot distinguish the two. Proteomics provides phenotypic validation and interpretation of genomics, enabling more precise and reliable information for early detection of liver disease^12^ and cardiovascular events^13^, classification of benign/malignant thyroid nodules^14^, and personalized prognostication^15^. Thus, proteomics drives the development of precision medicine. The above examples illustrate the promising prospects of proteomics technologies in the discovery of clinical biomarkers and drug targets, as well as disease classification.

In this study, we employed deep discovery and targeted proteomic strategies to select a panel of proteins and built a classifier for stratifying follicular neoplasms in retrospective tissue samples. The model was further validated in two independent retrospective tissue sets and prospective biopsy sets.

## Results

### Patient characteristics and study design

In the present study, we collected a total of 2443 data files which were analyzed for gene alterations and proteome profiling, from 1568 patients (including 909 with FTA and 659 with FTC) across 24 centers in China and Singapore. The median age at diagnosis was 49.0 years (interquartile range [IQR, Q1-Q3]: 36.0-60.0). There were 1105 females and 463 males, with a female-to-male ratio of 2.4:1. The median nodule size was 35 mm, with an IQR of 25.0 to 48.0 mm. Nodule sizes less than 40 mm were observed in 893 cases (57.0%), while sizes greater than or equal to 40 mm were observed in 667 cases (42.5%). Detailed patient information is listed in **Table 1**.

**Table 1.**
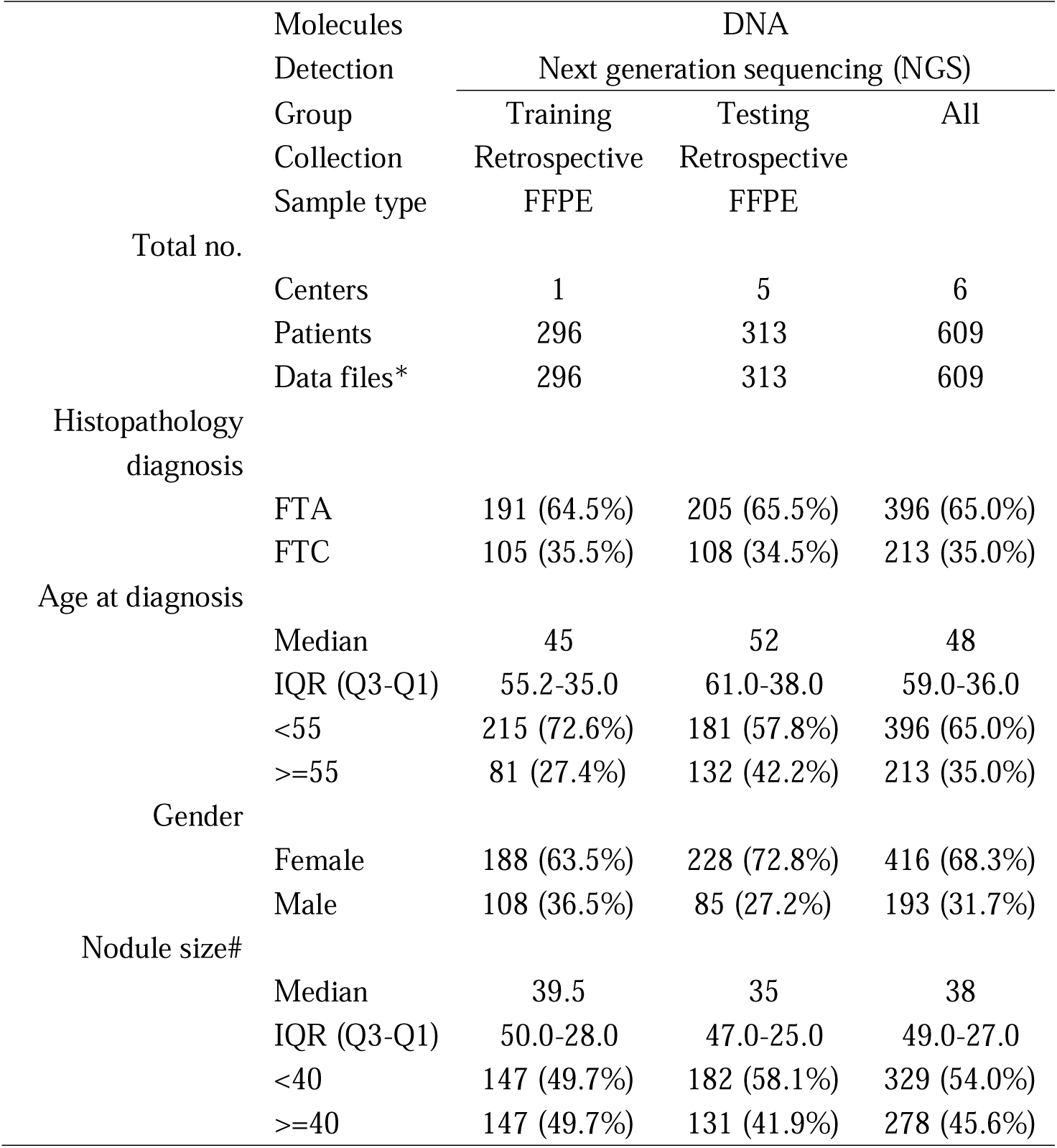
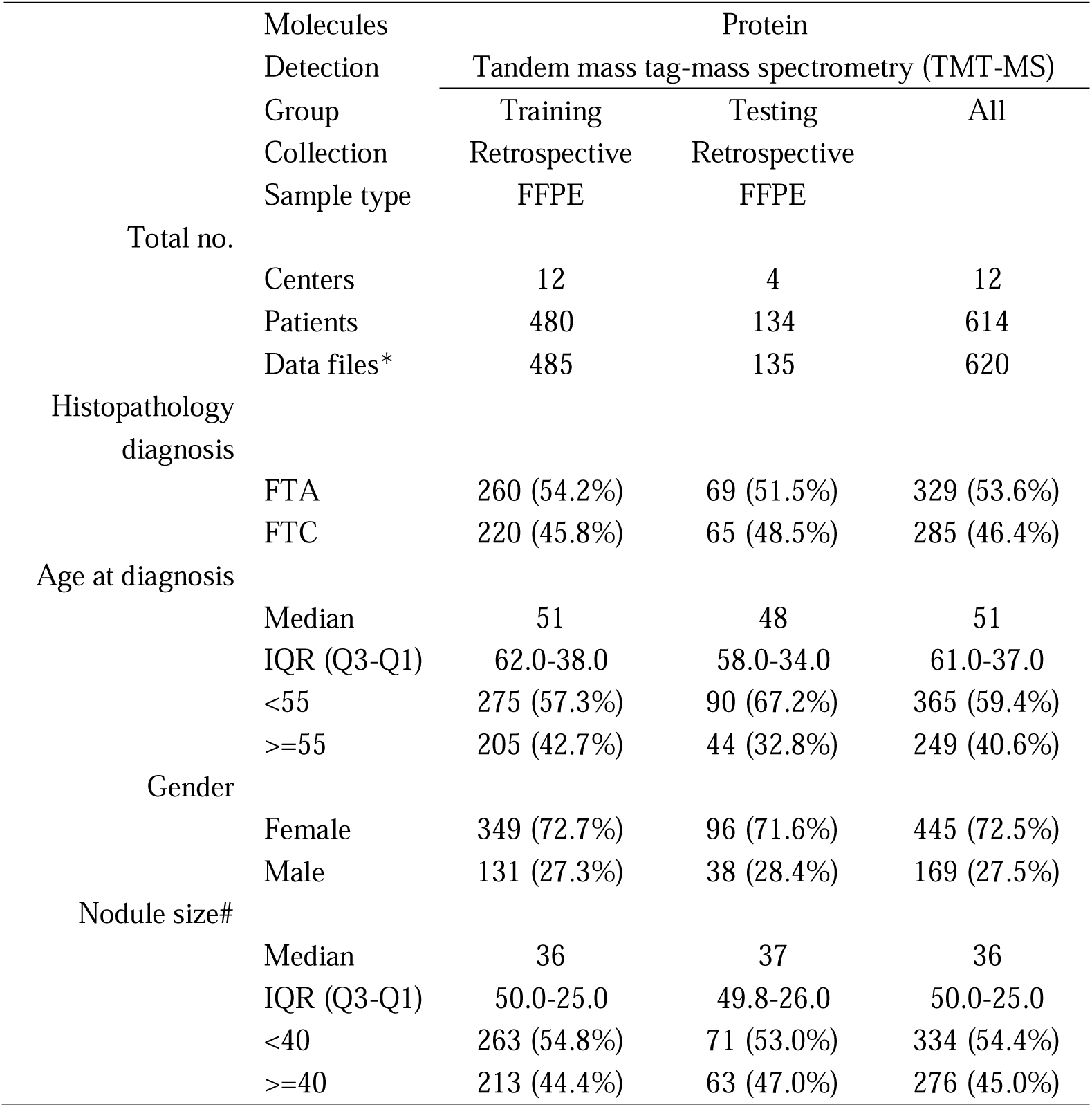
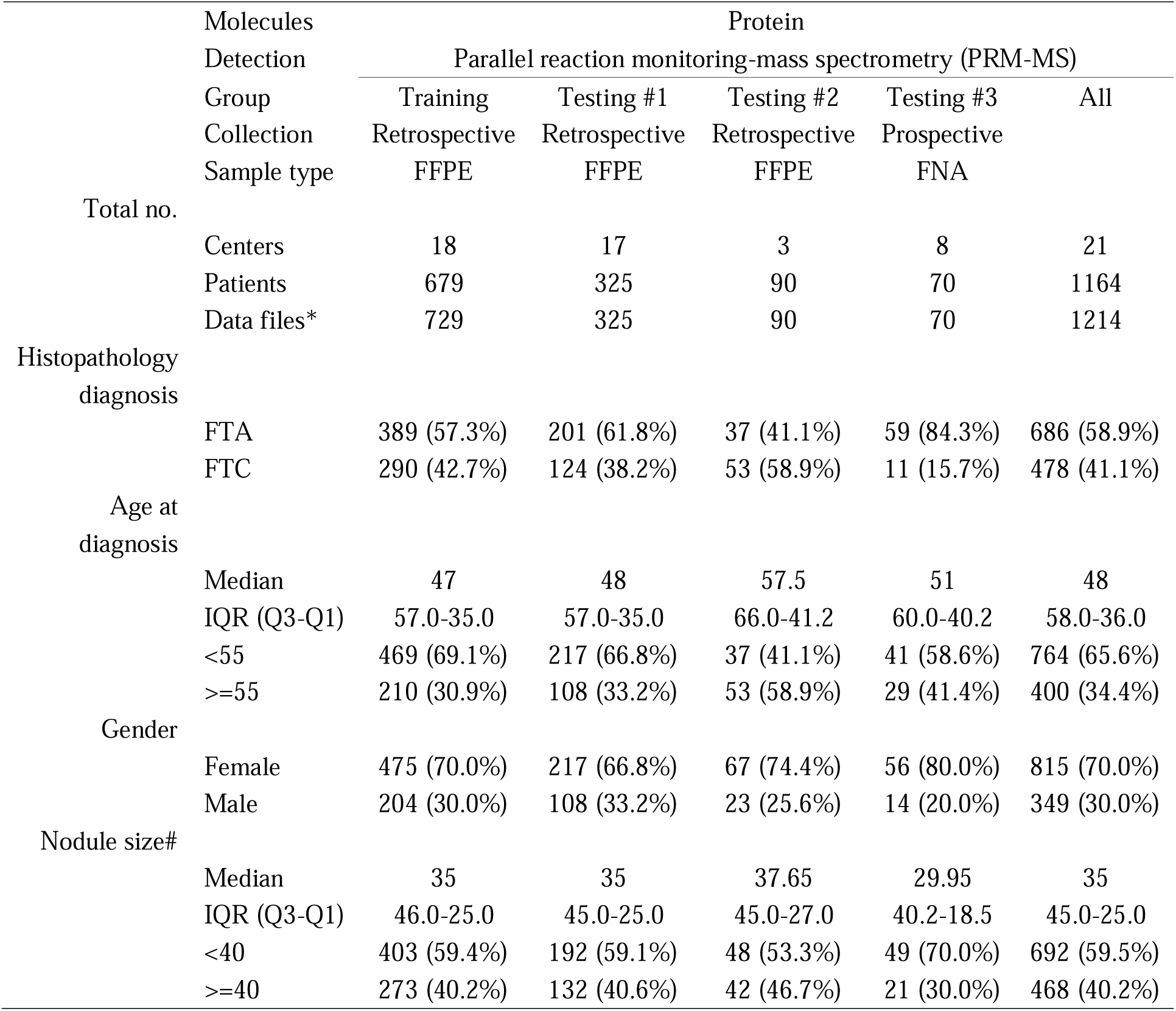
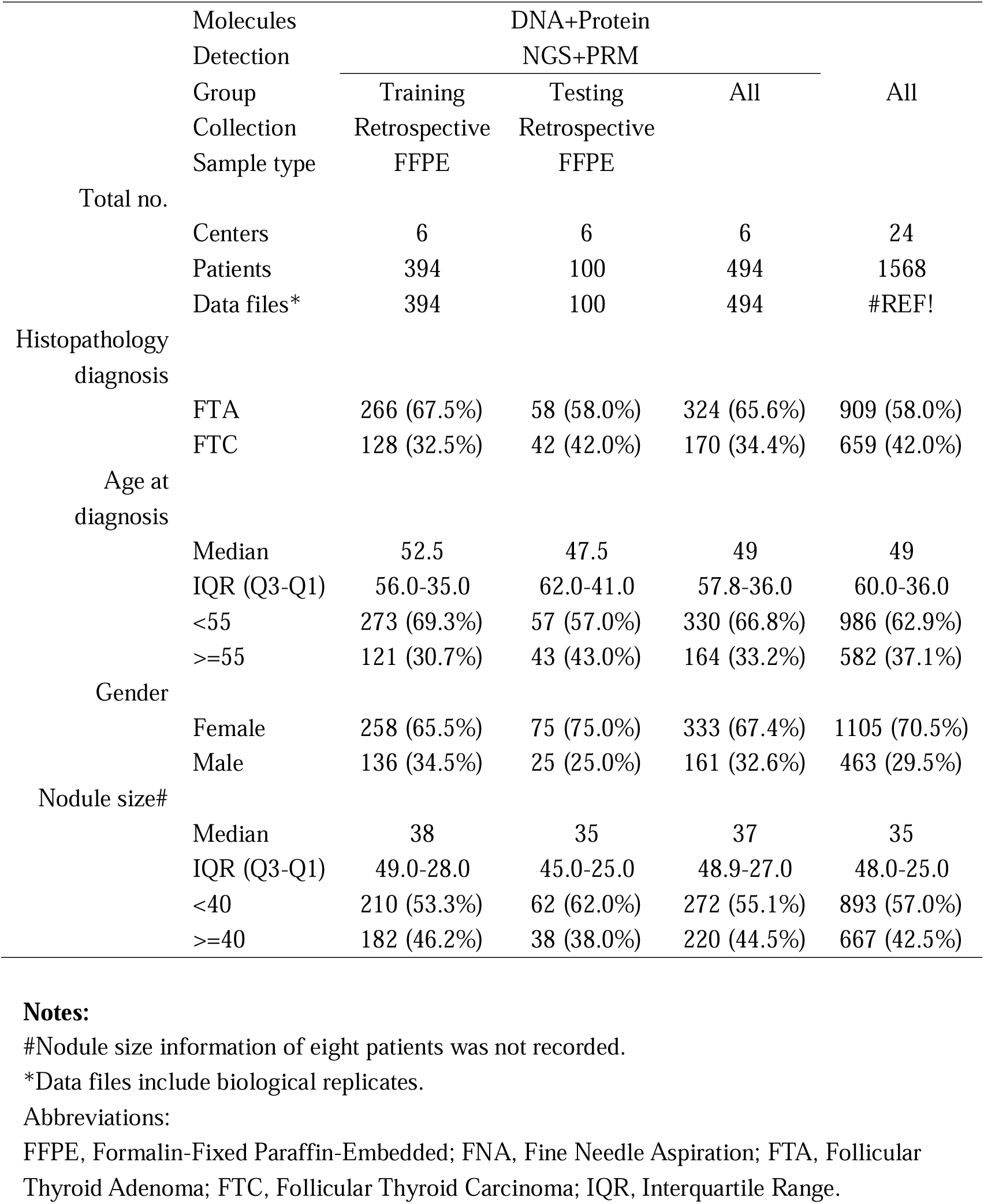
Baseline characteristics of patients from different sample

To profile the molecular landscape of follicular tumors, we first analyzed gene alterations of 609 samples from 609 patients by next-generation sequencing (NGS) and proteome differences between FTC and FTA of 620 samples from 614 patients by tandem mass tag (TMT)-based discovery proteomics. Next, we performed targeted proteomics through parallel reaction monitoring (PRM) on 729 samples to construct a classifier for stratifying FTC and FTA. The classifier was further validated in three testing sets including an internal testing set (n=325) and two independent retrospective (n=90) and prospective (n=70) testing sets (**Figure 1**). Finally, we compared the performance of combined gene panel data and targeted proteomics data from the same 494 patients against individual feature types to assess improved differentiation capability.

**Figure 1.**
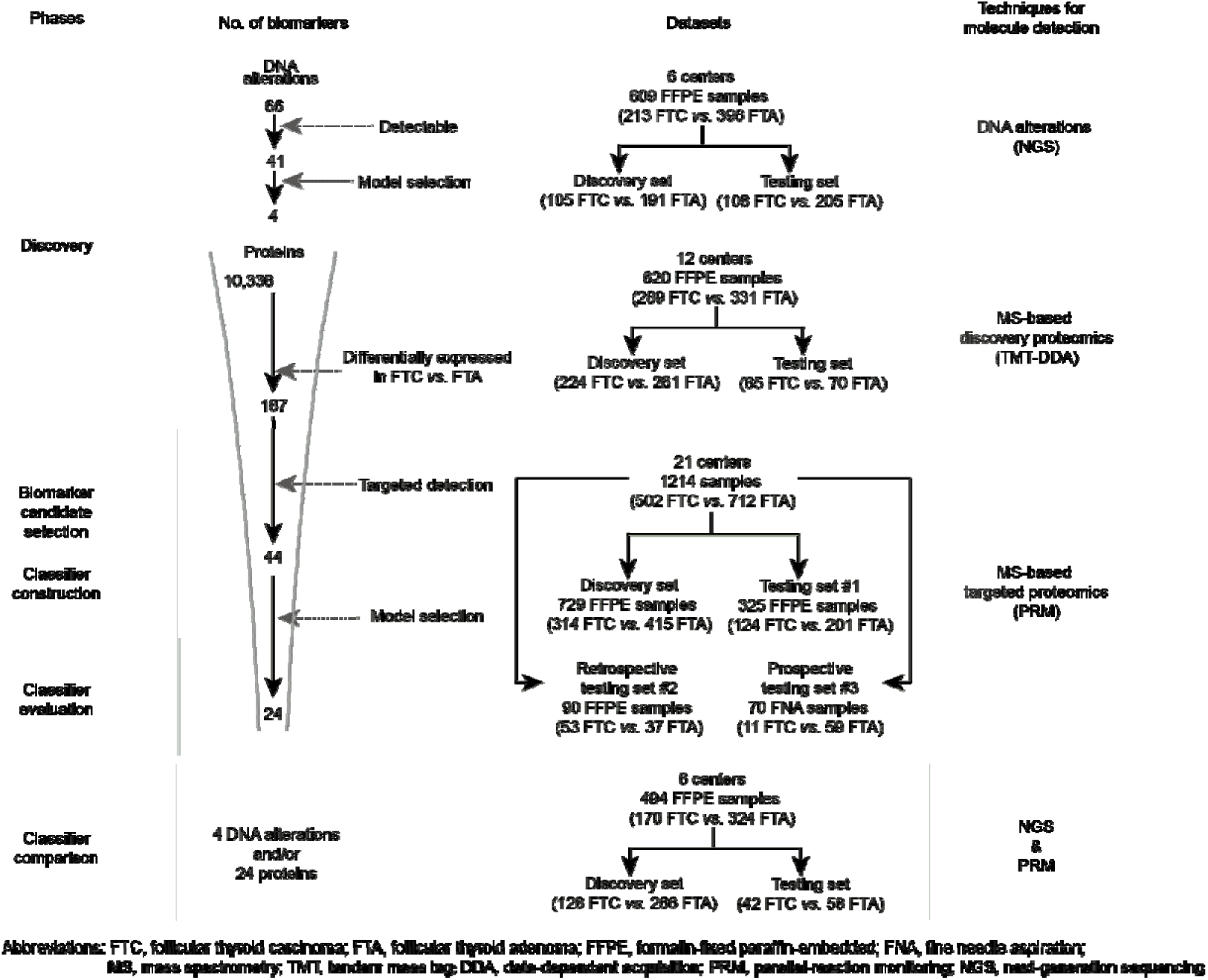
The flowchart of biomarker discovery, classifier development, performance evaluation, and comparison. The entire experiment was conducted in the following phases: discovery of dysregulated molecules, biomarker candidate selection, classifier construction, classifier evaluation, and comparison. The molecules and samples involved at different stages are illustrated in the corresponding phases.

### The gene-panel-based model cannot precisely distinguish FTA and FTC

We initially screened a 66-gene panel across 609 samples, including 396 FTA and 213 FTC. Among the 66 target genes, 41 genes (62.1%) were successfully detected in the present dataset, while the remaining genes were not. The gene analysis revealed that 325 of the 609 samples (53.4%) carried mutations, specifically 183/396 (46.2%) in FA and 142/213 (66.7%) in FTC. Moreover, 91 of 609 samples carried at least two mutations. The above data indicate that DNA alterations are uninformative of the histological diagnosis of 46.6% of follicular neoplasms.

The results exhibited a similar gene alteration pattern with different mutant frequency between FTA and FTC. The five most common mutations were *NRAS* (12.4% vs. 21.6%), *HRAS* (7.2% vs. 14.1%), *TERT* (2.3% vs. 18.8%), *DICER1* (4.0% vs. 10.3%), and *EIF1AX* (5.8% vs. 6.6%), with the numbers in parentheses indicating the mutation frequencies in FTA and FTC, respectively (**Supplementary Table 1**).

Subsequently, we constructed a gene-panel-based XGBoost model utilizing a matrix comprising 609 samples and 41 detected genes. The model was trained on a dataset comprising 296 samples from one center and subsequently tested on 313 samples from five independent centers. The model with four gene mutations (*TERT* promoter, *NRAS*, *DICER1* and *BRAF*) achieved an AUC of 0.670 (95% confidence interval [CI], 0.612-0.729), indicating that it was not sufficiently robust in the classification of FTA and FTC (**Figure 2a and Supplementary Figure 1**).

**Figure 2.**
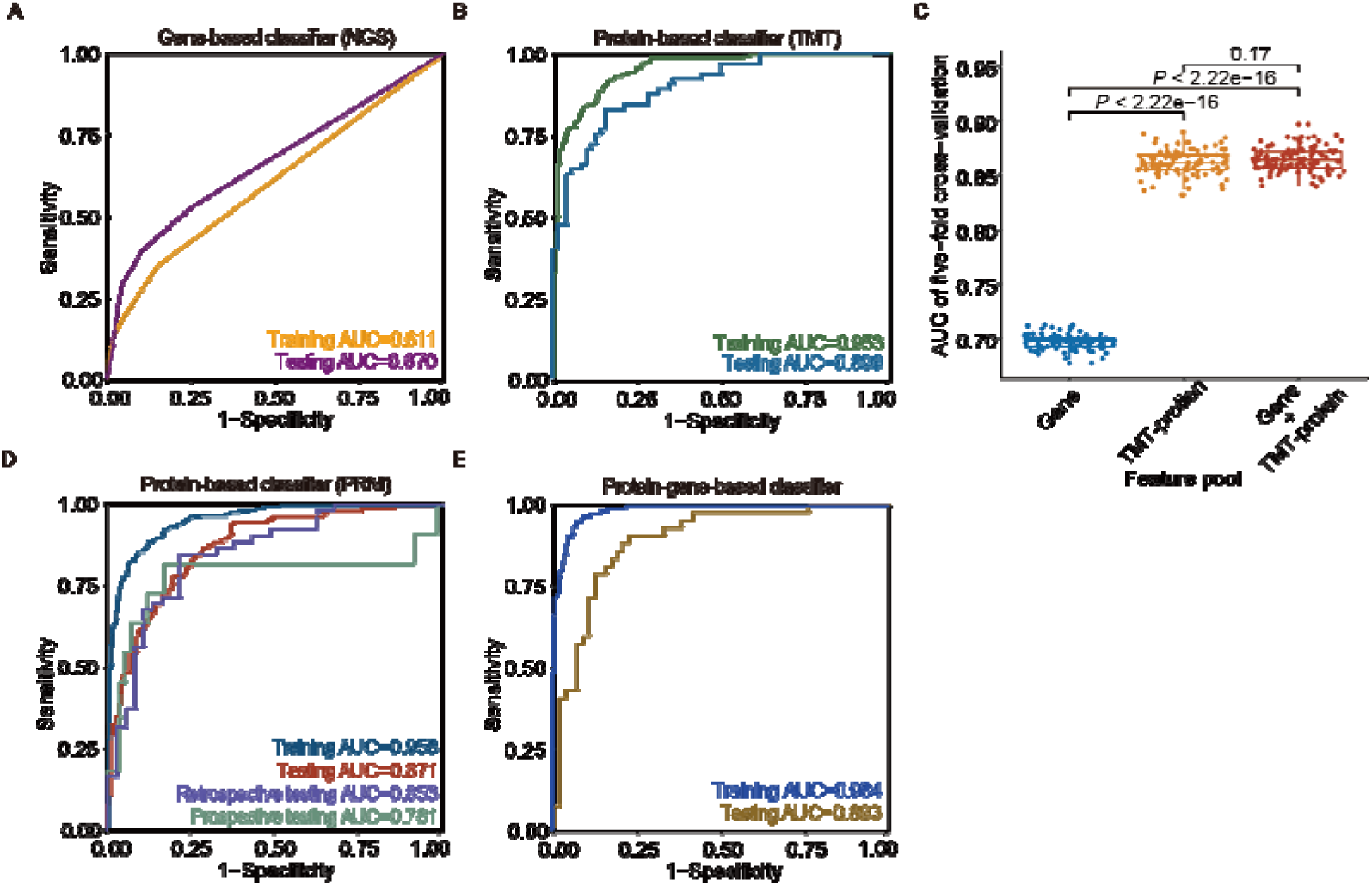
Performance of models across different datasets. Area Under the Curve (AUC) plots for (A) four-gene-based classifier detected by next-generation sequencing (NGS), (B) 24-protein-based classifier detected by tandem mass tag (TMT) discovery proteomics, (D) 24-protein-based classifier detected by parallel reaction monitoring (PRM) targeted proteomics in retrospective and prospective sample sets, (E) four gene and 24 protein combination-based classifier. (C) The AUC comparison of gene-, TMT-protein-, gene and TMT-protein-based models of five-fold cross-validation for one-hundred-time iterations. Each dot indicates one iteration. The boxes represent the first and third quartiles, the center line indicates the median, the whiskers extend to ±1.5 times the interquartile range, and the dots represent one iteration. *P* values are estimated by two-sided Welch’s *t*-test.

These findings suggest that gene mutations alone are insufficient to reliably distinguish between FTA and FTC which may be due to low-frequency gene alterations and their overlapping mutation patterns.

### In-depth proteomics analysis holds promise for differentiating FTA and FTC

Considering the similarities in biological morphology and gene expression between FTA and FTC, we employed an in-depth proteomics identification approach to analyze an FFPE dataset comprising 645 FFPE samples. From these samples, we quantified 10,336 proteins with a false discovery rate (FDR) of less than 1% at both peptide and protein levels. After quality control analysis (**Supplemental Figure 2**), we derived a matrix containing 7876 proteins from 620 samples (331 FTA and 289 FTC). The samples were further divided into a discovery set (485 samples, 261 FTA and 224 FTC) for protein feature selection and model construction, and an independent testing set (135 samples, 70 FTA and 65 FTC) for model performance evaluation.

Protein biomarker selection was conducted by comparison of FTC vs. FTA based on the discovery set which revealed 187 differentially expressed proteins (DEPs, **Supplemental Figure 3a**). Enrichment analysis showed these proteins to be involved in functions and pathways of thyroid hormone generation and metabolic processes (**Supplemental Figure 3b**). In dimensionality reduction analysis, we observed that the two groups of samples could be partially distinguished by DEPs, but the overall discriminability remains low, which is closely reflects the histological similarities between the two (**Supplemental Figure 3c**). Therefore, we further performed machine learning to improve discriminability.

After the performance comparison of six machine learning models with different feature counts (**Supplemental Figure 4**), the 24-protein-based XGBoost model was selected and applied in the following studies. Detailed model construction procedures and feature importance rank are shown in **Supplemental Figure 5**. Our model achieved AUC values of 0.953 (95% CI, 0.936-0.971), 0.905 (95% CI, 0.886-0.915) and 0.899 (95% CI, 0.849-0.949) in the training, cross-validation and independent testing sets, respectively (**Figure 2b**). In detail, the sensitivity, specificity, positive predictive values (PPV), negative predictive value (NPV), and accuracy were 0.800 (95% CI, 0.685-0.880), 0.843 (95% CI, 0.738-0.911), 0.825 (95% CI, 0.711-0.901), 0.819 (95% CI, 0.713-0.892), 0.822 (95% CI, 0.748-0.878), respectively, in the independent testing set (**Table 2** and **Supplemental Figure 5c**), which is much higher than the gene-based model. Next, we compared the model performance using gene-, TMT-protein- and combined feature pool. After 100 iterations of cross-validation testing, the model performance did not significantly improve with the addition of gene features on TMT-protein (**Figure 2c**), indicating that the additive role of genetic features is minimal. The above results illustrate the significant potential for discriminating FTA and FTC within retrospective samples. Therefore, in subsequent studies, we further developed clinically accessible protein-based classification measurements.

**Table 2.** Performance evaluation of three classifiers on the corresponding testing sets.

| Model | DNA-based | Protein-based (TMT) |
| --- | --- | --- |
| Feature no. | 4 | 24 |
| Datasets | Testing set | Testing set |
| Ratio of malignancy | 0.345 | 0.4815 |
| AUC | 0.670 (0.612-0.729) | 0.899 (0.849-0.949) |
| Accuracy | 0.719 (0.666-0.766) | 0.822 (0.748-0.878) |
| Sensitivity | 0.407 (0.320-0.502) | 0.800 (0.685-0.880) |
| Specificity | 0.883 (0.831-0.920) | 0.843 (0.738-0.911) |
| PPV | 0.647 (0.528-0.750) | 0.825 (0.711-0.901) |
| NPV | 0.739 (0.680-0.790) | 0.819 (0.713-0.892) |

**Table 2 continued**
| Model | Protein-based (PRM) |  |  | DNA&Protein -based |
| --- | --- | --- | --- | --- |
| Feature no. | 24 |  |  | 4+24 |
| Datasets | Testing set #1 | Testing set #2 | Testing set #3 | Testing set |
| Ratio of malignancy | 0.3815 | 0.5889 | 0.1571 | 0.42 |
| AUC | 0.871 (0.833-0.910) | 0.853 (0.772-0.934) | 0.781 (0.563-1.000) | 0.893 (0.829-0.957) |
| Accuracy | 0.785 (0.736-0.826) | 0.789 (0.692-0.861) | 0.757 (0.644-0.843) | 0.820 (0.732-0.883) |
| Sensitivity | 0.726 (0.641-0.797) | 0.792 (0.663-0.881) | 0.818 (0.510-0.957) | 0.714 (0.563-0.829) |
| Specificity | 0.821 (0.761-0.868) | 0.784 (0.625-0.888) | 0.746 (0.621-0.840) | 0.897 (0.788-0.954) |
| PPV | 0.714 (0.630-0.786) | 0.840 (0.711-0.918) | 0.375 (0.212-0.574) | 0.833 (0.676-0.924) |
| NPV | 0.829 (0.770-0.875) | 0.725 (0.570-0.839) | 0.957 (0.845-0.995) | 0.813 (0.698-0.890) |

**Notes:** Values (95% confidence interval) Abbreviations: AUC, Area Under the Curve; PPV, Positive Predictive Value; NPV, Negative Predictive Value.

### Targeted proteomics-based model development and evaluation

The previous in-depth proteomics approach is relatively costly and time-consuming for clinical diagnostic laboratories. In contrast, targeted proteomics, with its higher accuracy, stability, shorter run times, and cost-effectiveness, is more suitable for clinical application. Hence, we developed a targeted proteomic strategy for the protein biomarker candidates. A total of 44 proteins out of 187 DEPs were successfully detected using a single-injection targeted method. These proteins were further measured in four datasets (n=1214) comprising retrospective FFPE and prospective FNA samples from 21 clinical centers (**Figure 1**).

We initially analyzed 1054 samples from 18 centers, which were randomly allocated into discovery (n=729) and testing (n=325) sets. In the discovery set, the XGBoost algorithm was conducted and a panel of 24 protein biomarkers emerged and were ranked (**Supplemental Figure 6**). The characteristics of each selected protein with corresponding abundance are shown in **Supplemental Figure 7**. Of the 24 selected proteins, eight (CA4, ITIH5, FABP4, DPP4, CRABP1, HMGA2, TIMP1, ECM1) are known to be associated with follicular thyroid tumors, ten (MATN2, AHSG, CD36, STMN1, NPC2, IGF2BP2, P4HA2, LRP2, IGSF1, RAP1GAP) are related to other types of thyroid cancer, and the remaining six (CPOX, MYEF2, STEAP4, H1-5, FRAS1, TANC2) are newly discovered and have not been previously reported. After fine-tuning the hyperparameters, our model correctly identified 255 out of 325 samples with an accuracy of 0.785 (95% CI, 0.736-0.826), corresponding to a sensitivity of 0.726 (95% CI, 0.641-0.797) and a specificity of 0.821 (95% CI, 0.761-0.868). The PPV and NPV were 0.714 (95% CI, 0.630-0.786) and 0.829 (95% CI, 0.770-0.875), respectively, in the testing set (**Table 2**).

To further validate the generalization of our classifier, we additionally tested two independent sample sets, one of which was retrospectively acquired, and the other prospectively. In the retrospective testing set, the classifier accurately identified 78.9% of the samples corresponding to the sensitivity, specificity, PPV and NPV of 0.792 (95% CI, 0.663-0.881), 0.784 (95% CI, 0.625-0.888), 0.840 (95% CI, 0.711-0.918), 0.725 (95% CI, 0.570-0.839), respectively. We further tested the model on prospective FNA biopsies, which were obtained pre-surgery. The model also performed well, achieving a diagnostic accuracy of 0.757 (95% CI, 0.644-0.843) with sensitivity, specificity, PPV and NPV of 0.818 (95% CI, 0.510-0.957), 0.746 (95% CI, 0.621-0.840), 0.375 (95% CI, 0.212-0.574), 0.957 (95% CI, 0.845-0.995), respectively (**Table 2**).

Additionally, our model achieved AUC values of 0.871 (95% CI, 0.833-0.910), 0.853 (95% CI, 0.772-0.934) and 0.781 (95% CI, 0.563-1.000) in the cross-validation testing set, the independent retrospective and prospective testing sets, respectively (**Figure 2d**).

The foregoing multicenter testing results demonstrate that our protein-based model conducted by targeted proteomics can aid in the differential diagnosis of thyroid follicular tumors. It’s worth mentioning that this protein-based classifier enhances the accuracy of pre-surgical diagnoses for follicular thyroid tumors, which currently defies a definitive solution in clinical practice.

### Comparative evaluation of four genes and/or 24 protein-based models

To equally compare the models based on genes, proteins, and the combination of both, we utilized the same set of samples from the overlapping tissues in the two datasets (n=494) with the same data splitting. The hyperparameters of each model were optimized to ensure the best performance. Based on 5-fold cross-validation, the AUC of combined gene and protein features was significantly higher than only using 24 proteins (**Supplemental Figure 8a**). Moreover, the higher AUC owed more to protein features than to gene features (**Supplemental Figure 8b**). This model with combined features achieved an AUC of 0.893 (95% CI, 0.829-0.957) in the testing set (**Figure 2e)**. In detail (**Table 2** and **Supplemental Figure 8c**), the accuracy, sensitivity, specificity, positive predictive values (PPV) and negative predictive value (NPV) were 0.820 (95% CI, 0.732-0.883), 0.714 (95% CI, 0.563-0.829), 0.897 (95% CI, 0.788-0.954), 0.833 (95% CI, 0.676-0.924), 0.813 (95% CI, 0.698-0.890), respectively, in the independent testing set, which is comparable to the TMT-based model.

## Discussion

Although molecular tests have been adopted for the diagnosis of benign and malignant thyroid nodules^16^, differentiating FTC from FTA has long been a significant challenge in pathological diagnosis. This issue has persistently troubled clinicians and remains a particularly thorny problem. Despite numerous attempts over the years to distinguish between the two at various molecular levels, including DNA methylation^17,18^, mRNA^19–21^, DNA-mRNA^22^, and protein analyses^23^, no effective molecular markers or marker combinations have been identified to aid in clinical diagnosis.

Several factors may contribute to this limitation. First, there is insufficient detection depth. Since the boundary between FTA and FTC may not be well-defined and could represent different stages of the same disease, it is crucial to conduct deep, quantitative analyses to uncover subtle distinctions. Second, due to their inherent similarities, the identification of reliable molecular markers depends heavily on large-scale data to enhance statistical reliability. Multicenter studies that incorporate both retrospective and prospective approaches can further improve the robustness of biomarker discovery.

Therefore, we designed an international multicenter study to explore the potential strategy for discriminating FTA and FTC. Firstly, we assessed gene alterations in a large sample set containing 609 tissues the results of which were highly consistent with previous findings. Unlike high specificity (up to 98%) of *BRAF* V600E in papillary thyroid carcinoma^24,25^, no effective genetic markers have been identified to differentiate FTC and FTA, not even a panel of gene markers. Therefore, it is not surprising that our gene-based model could not distinguish the two. Proteins are the direct executors of biological activities, and some of them have been used as clinical biomarkers for various diseases, *e.g.* thyroglobulin (Tg) and thyroid peroxidase (TPO) for thyroid diseases, prostate-specific antigen (PSA) for prostate cancer, carcinoembryonic antigen (CEA) for liver cancer, and HER-2 for breast cancer subtyping. Based on these, we decided to compare the proteome of FTA and FTC through a deep proteome quantification strategy, namely TMT, for biomarker discovery and further developed a protein-based classifier by leveraging a clinically available targeted proteomic method, *i.e.*, PRM.

We quantified over 10,000 proteins by TMT, as a high-quality dataset in the field of proteomics, laying a solid foundation for subsequent biomarker discovery. From the deep quantification matrix, we identified 187 potential DEPs, a significant increase compared to the 14^26^ and 6^27^ DEPs identified in our previous studies. Next, 24 proteins filtered by XGBoost for high discriminant function achieved an AUC of 0.899. Compared to a gene-based model, a protein-based classifier has demonstrated significant potential for differentiation. Following this, we conducted targeted detection which can be adopted in the clinic. Targeted proteomics enables the rapid quantification of dozens to hundreds of proteins within a short time frame (typically from several minutes to tens of minutes), making it highly promising for clinical applications^28^. Similar methods are already in clinical use for detecting small molecules such as drugs, vitamins, and steroid hormones^29^. We next built a 24-protein-based model on a targeted proteomic matrix (44 proteins * 729 samples) and further validated it in two retrospective sets and a prospective testing set. The model achieved relatively high AUC of 0.871 and 0.853 in the retrospective sets. The lower AUC of 0.781 observed in FNA biopsies compared to FFPE samples can be attributed to the inherent sampling limitations and the heterogeneous nature of tumors. FNA biopsies, by their nature, sample only a small portion of the tumor, potentially missing areas of significant diagnostic or prognostic importance. Nevertheless, this protein-based classifier with a high NPV of 95.7% could improve diagnostic accuracy as a rule-out test to decrease unnecessary surgery on benign nodules.

The development of artificial intelligence offers more objective, accurate, and personalized options for medical evaluation^30,31^, especially for complex diseases. Due to inter-individual heterogeneity, relying on a single or a few molecular markers can be susceptible to noise interference, making it difficult to obtain reliable results. In contrast, using a panel of proteins (typically dozens) effectively mitigates this issue^32^. In this study, we balanced performance and the number of protein features, optimizing between 20-30 proteins, and ultimately identified 24 proteins to incorporate into the model. While proteins can be detected using antibody-based methods, clinical immunohistochemistry (IHC) typically allows for the detection of only one protein per assay, with the number of detectable proteins limited by the number of tissue sections. Although multi-color IHC is extensively used in scientific research, it has not yet been broadly adopted in real-world clinical diagnostics. Moreover, antibody-based quantitative detection is constrained by the specificity, sensitivity, and linear range of the antibodies, and is prone to be affected by experimental conditions and reagent batches. MS, on the other hand, offers high-throughput, high-precision, diverse, and consistent protein quantification^33,34^, making it more compatible with machine learning techniques to establish complex and reliable models for disease classification, diagnosis, and other applications. It is anticipated that MS will play an indispensable role in future clinical diagnostics and treatment.

Although we have achieved promising results, there are still some limitations that need to be addressed. First, there could be a sampling bias in this retrospective study, which may have affected the proposed test characteristics. Second, in the current model construction and evaluation, it is challenging to completely rule out false-negative samples, particularly those in the early stages of FTC without capsular invasion at the time of surgery. Third, the small number of patients of racial or ethnic minority groups included in this study may limit generalizability to these underserved groups.

In summary, this discovery investigation with subsequent prospective validation, shows that integrating deep proteomics and targeted proteomics coupled with machine learning facilitates precise diagnosis of follicular thyroid tumors. This paradigm can also be extended to the differential diagnosis of other types of diseases in the future.

## Materials and Methods

### Patients and samples

This study was approved by the Ethics Committee of Westlake University and each center. Our study sets were collected from 24 participating centers of the Westlake Thyroid Proteome Consortium (WE-TEC) working group. Clinical demographic data and histopathological reports were extracted from each medical record system. Histopathology was reviewed for hematoxylin and eosin (H&E) slides according to the standardized WHO classification (4^th^ and 5^th^ editions)^35,36^. Based on the pathological results, we included FTA and FTC samples and excluded samples with follicular variant papillary thyroid tumors.

In the retrospective sets, formalin-fixed paraffin-embedded (FFPE) slides or punches from 2002 to 2022 were collected. Additionally, we included 70 fine needle aspiration (FNA) biopsies with histopathology reports of follicular thyroid neoplasm from our registered prospective cohort from eight centers.

The sample datasets comprised four main parts. Firstly, there were 609 retrospective FFPE samples designated for DNA mutation detection. Secondly, 645 retrospective FFPE samples from 12 centers were used for protein biomarker discovery. Thirdly, 1054 retrospective FFPE samples were analyzed for targeted proteome analysis and classifier development. Lastly, the dataset included 90 retrospective FFPE samples and 70 prospective fine needle aspiration biopsies, which were utilized for independent model testing.

### Gene mutation analysis

Genomic DNA was extracted from thyroid FFPE slides utilizing the QIAamp DNA FFPE Advanced Kit (Cat. ID 56604, QIAGEN, Germany). Sequencing libraries were constructed using a designed thyroid cancer-related 66-gene panel (**Supplementary Table 3**), which is designed for multiplex targeted sequencing, enabling the detection of point mutations and insertions/deletions. An aliquot of extracted DNA was used for multiplex amplification of target regions, followed by polymerase chain reaction (PCR) amplification to add unique dual indices and Illumina sequencing adaptors (Illumina Inc., USA). Post amplification, the indexed libraries were purified with beads, quantified using a Qubit fluorometer (Thermo Fisher Scientific, USA), and sequenced to generate 150 bp paired-end reads on the NovaSeq 6000 platform (Illumina Inc., USA).

Quality assessment of the raw sequencing data was performed using FastQC (version 0.11.9). Adapter sequences and low-quality bases were trimmed from the raw reads using Cutadapt (version 1.18)^37^. The trimmed reads were then aligned to the human reference genome (hg19) with BWA (version 0.7.17)^38^. Variants, including single nucleotide variants (SNVs) and insertions/deletions (InDels), were identified using VarScan2 (version 2.4.4)^39^, and subsequently annotated using the Ensembl Variant Effect Predictor^40^.

### Sample processing for proteome analysis

FFPE tissue slides and FNA biopsies were processed using the pressure cycling technology (PCT)-assisted sample preparation pipeline as described in our previous publications^41,42^. Briefly, the FFPE samples were dewaxed with heptane and rehydrated using ethanol solutions of various concentrations. Following this, the samples underwent acidic hydrolysis with 0.1% formic acid and basic hydrolysis with 0.1 M Tris-HCl (pH 10.0). Proteins were then extracted using a lysis buffer containing 6 M urea and 2 M thiourea. The proteins were reduced and alkylated with 10 mM Tris (2-carboxyethyl) phosphine and 40 mM iodoacetamide under PCT assistance. Subsequently, PCT-assisted Lys-C/trypsin enzymatic digestion was performed, with the optimal enzyme-to-substrate ratios of 1:80 for Lys-C and 1:20 for trypsin.

TMT-based proteomics analysis was performed as previously described^43^. Briefly, 7 µg of peptides from each tissue sample and pooled peptide sample were labeled with the TMTpro 16-plex reagent (Thermo Fisher Scientific, San Jose, USA). Hydroxylamine was employed to quench the labelling reaction. Subsequently, the 16 TMT-labeled peptide samples were combined and cleaned using C18 columns. High-pH fractionation was then performed on a Thermo Ultimate Dionex 3000 (Thermo Fisher Scientific, San Jose, USA) equipped with an XBridge Peptide BEH C18 column (300Å, 5 µm, 4.6 mm × 250 mm) (Waters, Milford, MA, USA). The fractionation process utilized a 120-minute LC gradient, ranging from 5% to 35% acetonitrile (ACN) in 10 mM ammonia (pH 10.0), at a flow rate of 1 mL/min, resulting in 120 fractions. As previously described^43^, these 120 fractions were then combined into 30 fractions.

### Discovery proteomics data acquisition and processing

Peptides from each fraction were analyzed by an Orbitrap Exploris 480 mass spectrometry (Thermo Fisher Scientific, San Jose, USA) coupled with FAIMS Pro Duo interface (Thermo Fisher Scientific, San Jose, USA), along with a 60 min LC gradient at a flow rate of 300DnL/min. Eluted peptides Subsequently, the fractionated sample was separated with Thermo Scientific UltiMate 3000 RSLCnano System. The mass spectrometer was operated in positive mode with the FAIMS Pro interface and then analyzed with an Orbitrap Exploris 480 mass spectrometry by data-dependent acquisition (DDA) mode (Thermo Fisher Scientific™, San Jose, USA). The compensation voltage was set to -45V and -65V with a cycle time of 1 s per FAIMS experiment.

All the DDA data were processed using Proteome Discoverer version 2.4 (Thermo Scientific, USA) against a FASTA file downloaded from UniProt database (version 15/07/2020, 20368). The settings were referred to in a previously published paper. Briefly, missed cleavages within two were allowed. The minimal peptide length was set as six residues. Normalization was processed against the total peptide amount. Precursor ion mass tolerance was set to 10 ppm, and fragment mass tolerance was 0.02 Da. The false discovery rate (FDR) of peptides was set to 1% (strict) and 5% (relaxed).

### Data acquisition and processing of parallel-reaction monitoring (PRM)

PRM detection was performed preliminarily based on 187 selected proteins which were chosen from differentially expressed proteins (DEPs) in the TMT dataset and our previous published datasets^26,27^. After optimization, a set of 44 proteins from the original panel of 187 proteins. For each protein, we select one precursor to be monitored by Skyline (version 23.1). The selection criteria are as follows: a) no peptide modification, b) no missed cleavages, and c) peptide length ranging from 8 to 20.

Cleaned peptides were separated through UltiMate™ 3000 RSLCnano System (Thermo Fisher Scientific, San Jose, CA) equipped with a 15 cm x 75 µm analytical column (1.9 µm 100 Å C18-Aqua) through a 60-minute effective linear gradient of 6% to 30% buffer B (98% ACN, 0.1% formic acid) at 300 nL/min. The separated peptides were further analyzed by Q Exactive^TM^ HF (Thermo Fisher Scientific, San Jose, CA) with PRM data acquisition mode.

The time-scheduled acquisition mode was applied within a +/-3 min retention time window. The MS1 scans were collected *m/z* at 400 to 2500 Th with a resolution of 60,000 FWHM. The AGC target was set to 3E6 charges and the maximum IT was 55 ms. The target precursors were isolated through a window *m/z* of 1.6 Th. The normalized collision energy for fragmentation was set at 27%. The products were scanned at a resolution of 30,000 FWHM, the AGC target value was set to 5E5 charges, and the maximum injection time was 120 ms. Here, a total of 44 precursors from 44 proteins and 20 CiRT were analyzed. PRM data were further analyzed by Skyline (version 23.1) with the same setting when we developed the PRM method. Next, the protein abundance matrix was transformed by log2.

### TMT data quality control and preprocessing

We firstly excluded ten samples, of which two samples are not follicular tumors, six samples are from metastatic sites, and two samples have incomplete clinical information. For the pooled samples, data quality was assessed by analysis of the coefficients of variation (CV) across pooled samples. Next, principal component analysis (PCA) was performed based on the discovery set with missing values imputed with zero. We next deleted fifteen high-missing-proportion samples which were detected as outliers in the PCA. We ultimately analyzed a total of 620 FFPE tissue slides comprising 331 FA and 289 FTC from 12 clinical centers. Additionally, the proteins with NA rate > 60% were removed.

Missing values imputation was conducted by R package *NAguideR* ^44^ and the impseqrob algorithm was used. To avoid leakage of testing data, we imputed the discovery set first, which ensured the imputation for testing data did not affect the discovery set. Then, the testing set was combined with the filled discovery set and imputed by the same imputation method impseqrob.

Batch effects were detected by two-dimensional uniform manifold approximation and projection (UMAP) visualization. Batch effects were corrected by the Combat algorithm in R package *sva* ^45^, which is an empirical Bayes framework for adjusting data for batch effects. Similarly, we adjusted the batch effects of the discovery set first. As for the testing set, to prevent testing data or label leakage, we held the testing set label unknown for Combat and corrected batch effects referring to the discovery set after correction, keeping the discovery set unchanged.

### PRM data preprocessing

For PRM data, the Deterministic Minimal Value (MinDet) algorithm was first utilized for imputing the missing values in the matrix. For the retrospective and prospective testing sets, the ComBat algorithm was selected to deal with the batch effects, with the PRM training set as the reference dataset.

### Modelling

#### Gene-mutation-based modeling

The establishment of a gene-mutation-based classifier was based on the dataset of 609 patients from six centers. Patients from one center were selected as training sets and applied for model building. Samples from the other five centers were adopted for independent testing. Gene data in the analyzed matrix were transformed to logical data where 0 and 1 represent wide type and mutation, respectively.

#### TMT-based protein modeling

To build and evaluate our classification system, we split our data into two parts: a discovery set (n=485) and an independent testing set (n=135).

The machine learning-related contents were implemented by R package *mlr3*, which is a modern object-oriented machine learning framework in R. Before building machine learning models, to simplify the feature selection and engineering, we chose to narrow down the feature set by ensemble filtering. In detail, three filtering criteria, ANOVA *P* value, Kruskal-Wallis test *P* value and information gain were considered and features with ANOVA *P* values no less than 0.001 or Kruskal-Wallis test *P* values no less than 0.001 or no information gain would be removed.

Six machine learning algorithms^46^, K-Nearest Neighbor (KNN), Naïve Bayes (NB), Logistic regression (L), Support Vector Machine with radial basis function kernel (SVM), Random Forest (RF), and eXtreme Gradient boosting (XG or XGboost) were chosen as potential algorithms to deal with the FA and FTC classification task. Before benchmarking these algorithms, we tuned the hyper-parameters of the 6 machine learning models by random search (100 hyper-parameters combinations tried) and 5-fold cross-validation on the discovery set and determined the best hyper-parameter setting. Then to compare the models under the best hyper-parameter setting, we ranked the features by averaging the rankings of three filter criteria, used different feature number settings and conducted 100 times 5-fold cross-validation for each model under each feature number setting with AUC recorded. The best model was determined by referring to the average AUCs under different feature numbers and would be used for downstream analysis.

To refine the selected model, a more concise and model-based feature selection was conducted. In detail, we first trained XGboost on a discovery set 100 times with different random seeds, each time selecting the 50 most important features and retaining the features that appeared no less than 30 times. Additionally, the feature importance mentioned above was calculated by gain which represents the fractional contribution of each feature to the model based on the total gain of this feature’s splits and a higher percentage means a more important predictive feature.

#### PRM-based protein modeling

For PRM modeling, the proteins with fold change > 1.4 and Benjamini & Hochberg (BH)-adjusted Welch’s t-test P value < 0.05 were selected as important features for further modeling. For XGBoost model building, the hyperparameters (lambda, alpha and nrounds) were tuned by 5-fold cross-validation and random search based on the training set, also towards the highest classification AUC.

#### Modeling based on gene and PRM-based protein data

For Gene and PRM modeling, firstly, we only retained the four feature genes and 24 feature proteins that were selected by previous models. The applied dataset (n=494) contains both gene-mutation and PRM-based protein data. The training set (n=394) for this comparison was derived from the previously established model training set combination, and the remaining samples were designated as the testing set (n=100). Next, we conducted 100 times 5-fold cross-validation to compare gene-based, PRM-based and gene & PRM-based modeling. For XGBoost model building, the hyperparameters (lambda, alpha, and nrounds) were tuned by 5-fold cross-validation and random search based on the training set, also towards the highest classification AUC.

### Statistical analysis

The statistical and bioinformatic analyses were conducted by R (version 4.2.3). PCA in the quality control part was performed based on a centered but not scaled discovery set with missing values imputed by zero. UMAP was implemented by function umap in R package *umap* with default parameter setting. The Welch’s *t*-test was utilized to compare the expression difference of proteins between FA and FTC and the resulting p-values were adjusted by the BH method. The confidence intervals for training and testing performance measures except for AUC were Wilson’s 95% confidence interval, and the variance of the AUC was computed as defined by the bootstrap method by the pROC package and the 95% confidence interval was deduced with the normal distribution. As for the confidence intervals of cross-validation performance measures, they were approximately 95% confidence intervals from mean - 1.96*standard deviation to mean + 1.96*standard deviation. The cross-validation ROC originated from averaging five ROC curves in 5-fold cross-validation.

## Data Availability

All the proteomic raw data have been deposited to the ProteomeXchange Consortium (http://proteomecentral.proteomexchange.org) under the identifier IPX0008384000. Specifically, the TMT, PRM discovery, and PRM testing raw files are available under the identifiers IPX0008384001, IPX0008384003, and IPX0008384002, respectively.

All the data will be made publicly available upon publication of this manuscript. Please access the data via the following link before the official publication of the paper.

## Code Availability

The code for statistical analysis and modeling presented in this manuscript and generating corresponding figure panels and tables are publicly available on GitHub.

## Acknowledgments

This work was supported by the National Key R&D Program of China (Grant No. 2022YFF0608403, 2021YFA1301600), and the China Postdoctoral Science Foundation (2022M722841), the National Natural Science Foundation of China (Key Joint Research Program) (Grant No. U21A20427). The project was also supported by the Pioneer and Leading Goose R&D Program of Zhejiang (2024SSYS0035). We thank the Westlake University Supercomputer Centre for data storage and computation. Gene alteration detection was supported by RIGEN Biotechnology Co., Ltd, special appreciation goes to Dr. Wentian He and Dr. Yuxin Li for their valuable insights on gene analysis.

## Author Contributions

T.Guo., H.G., and Y.Zhu. supervise for this project. Y.S. and T.Guo. conceived and designed the study. Jianbiao.W., W.C., L.P., W.S., Z.W., Y.H., G.W., Y.Zhao., Y.D., W.W., F.W., D.L., X.L., H.Zheng., X.Z., B.W., J.S., J.Yao., Z.Yuan., T.L., J.P., Y.Zhang., Y.L., Q.G., Q.W., T.Gong, T.C., S.Z., J.Z., H.L., C.C., H.H., S.S., Y.Wu, Y.Wang, D.X., H.Zhang., X.N., N.G.I., Z.L. and H.G. provided tissues and patient information. F.Y. and H.G. provided gene alteration information. Y.S., L.L., P.H., J.Yu., N.Yao., Jiatong.W. and Z.G. participated in proteomic sample processing and data acquisition. Y.S., H.W., L.L., Y.Z., X.C. and W.G. conducted data analysis. All authors were responsible for the acquisition, analysis, or interpretation and discussion of data. Y.S., H.W., and L.L. drafted the manuscript. All authors provided critical revision of the manuscript for important intellectual content.

## The whole list of We-TEC investigators

Bei Wei^19, 20^, Bo Huang, Chuang Chen^30^, Deguang Zhang, Dingcun Luo^17^, Dong Xu^19, 20^, Erik K. Alexander^35^, Fan Wu^17^, Feng Yang^9^, Gaosong Wu, Guan Ruan, Guang Chen, Guangzhi Wang^13^, Guoyang Wu, Haitao Zheng^18^, Haixia Guan^39^*, Hanqing Liu^30^, Hao Zhang^10^, He Wang^1, 2, 3#^, Hong Han^31^, Huanjie Chen^14^, Huixiong Xu, James A. Fagin, Jiafei Shen^19, 20^, Jianbiao Wang^5#^, Jiang Zhu, Jianhua Wang, Jiatong Wang^1, 2, 3^, Jincao Yao^19, 20^, Jing Liu, Jingqiang Zhu^29^, Jun Pan^23^, Kennichi Kakudo^34^, Lei Liang, Lei Xie, Li Peng^7#^, Lu Li^1, 2, 3, 4#^, Meiping Shen, Mengni Chen^8^, Mingzhao Xing^33^, N. Gopalakrishna Iyer^37^, Oi Lian Kon^36^, Pingping Hu^1, 2, 3^, Qiaonan Guo^25^, Qijun Wu^26^, Qiushi Zhang, Sathiyamoorthy Selvarajan^32^, Shu Zheng^28^, Tiannan Guo^1, 2, 3, 40, 41^*, Ting Chen^28^, Tingting Gong^27^, Tong Liu^22^, Wanyuan Chen^6#^, Wei Sun^10^, Weigang Ge^8^, Wen Tian, Wenjun Wei^16^, Xiangfeng Lin^18^, Xianghui He, Xiao Shi, Xiaodong Teng, Xiaohong Wu^15^, Xin Zhu^19^, Xiu Nie^7^, Xue Cai^1, 2, 3^, Yan Li, Yan Zhou^1, 2, 3^, Yangfan Lv^25^, Yaoting Sun^1, 2, 3#^, Yi He^12^, Yi Zhu^1, 2, 3^*, Yifeng Zhang^24^, Yijun Wu^23^, Yingrui Wang^1, 2, 3^, Yongfu Zhao^13^, Yu Wang^16^, Yuxin Du^16^, Zhennan Yuan^21^, Zhihong Wang^10^, Zhiqiang Gui^10^, Zhiyan Liu^38^ The contributing We-TEC investigators are listed in the author list with affiliations. The non-contributing We-TEC investigators of this project are listed in the supplementary file.

## Supplemental Figures and Tables

**Supplemental Figure 1. Features and evaluation result of gene-based model**. (A) The importance rank of four selected features. (B) Confusion matrix of the four-gene model.

**Supplemental Figure 2. Discovery proteomics design and quality control. Study flowchart.** Samples (n=620) were retrospectively collected from 12 clinical centers. The samples were randomly allocated into 43 batches and prepared by pressure cycling technology (PCT) assisted sample preparation. Peptides were further labeled by 16-plex tandem mass tag (TMT) and data were acquired through the mass spectrometer with data-dependent acquisition (DDA) mode. There were 10,336 proteins quantified, and a 24-protein classifier was built based on them. (B) Uniform manifold approximation and projection (UMAP) plot showing the distribution of batches of samples using 7876 proteins. Dots are colored according to the TMT batches. (C) Violin plots show the CV of protein abundance across pooled samples in the discovery and testing sets. Medians are labeled upon the plot. The boxes represent the first and third quartiles, with the center line indicating the median. The whiskers extend to ±1.5 times the interquartile range, while the violin shapes depict the density of the data points.

**Supplemental Figure 3. Comparative proteomic analysis for FTC vs. FTA in TMT discovery dataset.** (A) Differentially expressed proteins (DEPs) in FTC and FTA. Thresholds of significantly dysregulated proteins: fold change>1.2 with Benjamini & Hochberg adjusted *P*<0.05 (Welch’s *t*-test). The highlighted proteins are the features selected by the model in **Supplemental Figure 4**. (B) Gene ontology (GO) biological process enrichment. X-axis represents the *P* and node size indicates the protein count of enriched items. *P* values are calculated using a one-sided Fisher’s Exact Test. (C) UMAP plot showing FA and FTC are partially resolved using 187 DEPs.

**Supplemental Figure 4. Comparison of machine learning models and feature counts performances.** (A)The compared six models (x-axis) are k-nearest neighbors (KNN), naive bayes (NB), linear regression (LR), support vector machine (SVM), random forest (RF), extreme gradient boosting (XGBoost). Each panel indicates the different protein feature counts applied. Each boxplot was derived from 100 times cross-validation on the training set. Y-axis showing the values of area under the curve (AUC) from the 100 iterations. The boxes represent the first and third quartiles, the center line indicates the median, the whiskers extend to ±1.5 times the interquartile range, and the dots represent individual data points.

**Supplemental Figure 5. Modelling using the discovery proteomic data.** (A) Schematic of XGBoost model construction. Samples firstly are divided into a discovery set and an independent set. Feature selection and model training are based on the discovery set and model performance evaluation is based on the independent test set. (B) Importance ranking of selected protein features and the frequency of 24 feature proteins when conducting 100 times feature selection on the training set. The orange color bar shows the selection frequency. (C) Confusion matrix of the 24-protein classifier. The blue color bar indicates the sample counts.

**Supplemental Figure 6. Targeted proteomic data-based feature importance and model performance.** (A) The 24 selected targeted protein features and their importance rankings. (B) Confusion matrix of the 24-protein classifier. Colors indicate the sample counts.

**Supplemental Figure 7. Protein feature characteristics.** (A) The protein identified by Uniprot ID_protein name_gene name with corresponding sequences which detected by targeted proteomics. (B) A chromatographic profile of a representative peptide precursor peak group. (C) Boxplots showing the protein abundance of each sample measured by parallel-reaction monitoring. FTA (n=415) and FTC (n=314). The boxes represent the first and third quartiles, the center line indicates the median, the whiskers extend to ±1.5 times the interquartile range, and the dots represent individual data points. *P* values are calculated by two-sided Welch’s *t*-test.

**Supplemental Figure 8. Model performance comparison and characters of the combined feature-based model.** (A) The area under the curve (AUC) comparison of gene-, protein-, gene and protein-based models of five-fold cross-validation for one-hundred-time iterations. Each dot indicates one iteration. The boxes represent the first and third quartiles, the center line indicates the median, the whiskers extend to ±1.5 times the interquartile range, and the dots represent one iteration. *P* values are estimated by two-sided Welch’s *t*-test. (B) The importance ranking of features. Protein features are colored in blue and gene features are colored in red. (C) Confusion matrix of the combined feature classifier. Colors indicate the sample counts.

**Supplemental Table 1. Gene mutation frequency in FTA and FTC.**

**Supplemental Table 2. Twenty-four proteins established associations with thyroid physiology or pathology.**

**Supplemental Table 3. List of 66-gene panel of the thyroid cancer.**

